# Hepatitis B cell-free DNA in non-invasive prenatal testing as an early biomarker of viral infectivity

**DOI:** 10.64898/2026.08.15.26360031

**Authors:** Vinh Nguyen Dao, Phuc Tri Nguyen, Thang Nhat Tran, Sang Hung Tang, Hoai-Nghia Nguyen, Maciej F. Boni, Hoa Giang, Minh-Duy Phan

## Abstract

Non-invasive prenatal testing (NIPT) was initially developed to detect chromosomal abnormalities in fetuses through the analysis of cell-free fetal DNA in maternal blood. Recent advancements have expanded NIPT’s applications to include the detection of viral infections during pregnancy. However, interpreting pathogen-derived cell-free DNA (cf-DNA) remains clinically complex. This study explores the clinical relevance of hepatitis B virus (HBV) cf-DNA using a dataset of approximately 500,000 NIPT visits and an independent validation cohort of 582 pregnant women (40 HBV-infected), aligned with HBV epidemiology from both population and individual perspectives. Our analysis reveals that HBV cf-DNA is a strong biomarker of high viral infectivity rather than a general marker of infection, suggesting its potential to identify pregnant women at heightened risk of vertical transmission by the end of the first trimester. Additionally, HBV-positive women showed a small but consistent reduction in fetal fraction relative to HBV-negative women across gestational weeks 9–17, an association compatible with an early effect of HBV on the placental contribution to cell-free DNA, although the observational design and unmeasured maternal covariates preclude causal inference.

## Introduction

Hepatitis B virus (HBV) infection continues to be a significant global public health challenge, particularly in regions with high endemicity. Among pregnant women, HBV presents a dual threat, impacting maternal health and posing a substantial risk of vertical transmission to the newborn. This mode of transmission is particularly concerning, as it can result in chronic HBV infection in approximately 90% of infected infants ^1,2^ ^3–5^, leading to long-term complications such as hepatocellular carcinoma and liver cirrhosis. Despite the availability of HBV vaccination and other prevention strategies, mother-to-child transmission (MTCT) remains a critical concern ^6–10^, particularly in low-resource settings where access to comprehensive care may be limited.

Vietnam, which is one of the ten countries contributing to two-thirds of the global HBV burden ^11^, reflects the broader HBV epidemiological trends observed across Southeast Asia ^12^. The prevalence of chronic HBV infection in Vietnam is estimated to range between 8% and 20% in the general population ^13–18^, with approximately 10% of pregnant women infected ^19,20^. This elevated prevalence positions HBV as one of the most critical infections during pregnancy in Vietnam. Effective prevention of mother-to-child transmission (MTCT) of HBV in high-endemicity countries hinges on two primary interventions: maternal antiviral therapy during pregnancy and the timely administration of the HBV vaccine along with hepatitis B immunoglobulin (HBIG) to the newborn within 24 hours of birth ^21^. Despite the established role of maternal viral load as a key biomarker for the risk of vertical transmission ^9,10,22–28^, this factor is frequently overlooked in pregnant women, with a greater emphasis placed on infection status alone. Consequently, many pregnant women in high-endemicity countries initiate antiviral therapy late, and some opt out of treatment altogether due to the limited remaining time to delivery. This highlights the critical need for early stratification of HBV infectivity and timely therapeutic intervention during pregnancy to improve management and reduce risk of MTCT.

Non-invasive prenatal testing (NIPT) was first introduced in 2011 as a method for detecting chromosomal abnormalities in a fetus using cell-free fetal DNA circulating in the mother’s blood ^29–32^. Recent advances have expanded the application of NIPT to detect viral infections during pregnancy ^33–35^. By analyzing the maternal blood samples, healthcare providers can monitor the presence of viral DNA, which provides crucial information for managing and mitigating risks to both the mother and the developing fetus.

This study represents a pioneering approach in assessing the potential of non-invasive prenatal testing (NIPT) for the early detection of hepatitis B virus (HBV) infection in the late first trimester. Here, we present an analysis that integrates our NIPT dataset with HBV epidemiological data at both the population and individual levels. We then emphasize the clinical significance of NIPT in identifying the risk of vertical transmission early in pregnancy.

## Materials and Methods

### Participants

A total of 499,544 pregnant Vietnamese women who underwent non-invasive prenatal testing (NIPT) between 2016 and 2024 at the Medical Genetics Institute in Vietnam were included in this study. The dataset was previously processed and published ^36^. The dataset was divided into two cohorts: 495,603 women who underwent a single NIPT visit (SD) and 3,941 women with at least two NIPT visits due to multiple pregnancies (DD). Data from third or subsequent visits were excluded. To validate our findings, we compared the NIPT results with the Hepatitis B e Antigen (HBeAg) and HBV-DNA tests, which are key biomarkers for assessing active viral replication and viral load. We conducted a retrospective analysis of 582 pregnant women who underwent NIPT tests at the University Medical Center, Ho Chi Minh City. Among those, there are 40 infected women (positive under Hepatitis B Surface Antigen (HBsAg) test). This small validation cohort included 12 women and 28 women who are HBV positive and negative under NIPT results respectively. The retrospective analysis was approved by the Institutional Ethics Committee of the University of Medicine and Pharmacy, Ho Chi Minh City, Vietnam.

### Combined HBeAg/HBV-DNA outcome

It is important to notice that not all 40 pregnant women observed at BVDHYD underwent both HBeAg and HBV-DNA testing simultaneously. At some analyses, we integrated the results of these two tests into a unified outcome measure, termed the “combined outcome” or “combined test.” A woman was considered positive at any given time point under the combined test if either the HBeAg or HBV-DNA result was positive. For instance, if HBV-DNA levels exceeded 200,000 IU/mL—the threshold for recommending HBV treatment — she was classified as positive. Conversely, if HBV-DNA levels were below this threshold and HBeAg was negative (if applicable), she was categorized as negative under the combined test.

### Age-specific Prevalence Modeling

#### Single NIPT visit data

Let *p_a_* denote the prevalence of hepatitis B virus (HBV) among women in age group *a*. The parameter *m* represents the probability that an infected woman will test positive under the NIPT method. Consequently, 1 − *m* indicates the probability of a false negative. It is assumed that uninfected women will always be correctly identified as negative with a probability of 100%. For a woman with one NIPT visit, the probability of observing negative (neg) or positive (pos) status is given as:

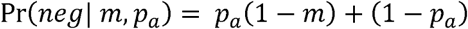

and:

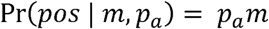

The likelihood of observing the single NIPT dataset is defined as the product of observing each data point:

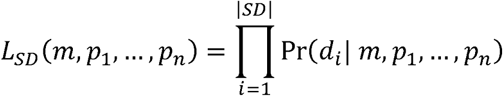

Where *d_i_* is the test result of the woman *i* in *SD*. The explanation in detail for each situation can be seen in Figure S1.

#### Double NIPT visit data

For a woman *i* in the double visit dataset, let *t_i_* represent the interval between her two visits. If a woman is uninfected at the first visit, she is assumed to be subject to a force of infection *ft_i_* during the interval. Conversely, if she is infected at the initial visit, she is assumed to undergo viral clearance at a rate of *μ* of *μt_i_*. Given that the force of HBV infection and HBV clearance rate are typically low in adults, we assume that the infection status of any pregnant woman is likely to change at most once between the two visits. Given these assumptions, the probability of observing a woman who tests positive at both visits is:

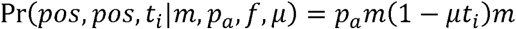

For the scenario where two tests are not consistent, the probability is given by:

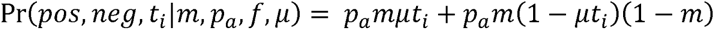

and:

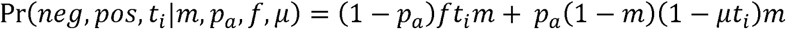

For the scenario where a woman tests negative at both visits, her true infection status could vary. Thus, the probability is determined by:

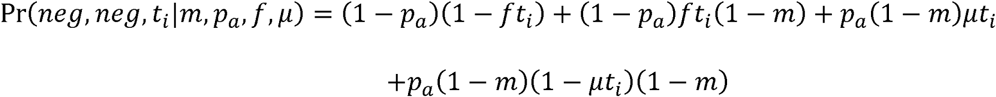

The likelihood of observing the double visit data set is defined as the product of observing each data point:

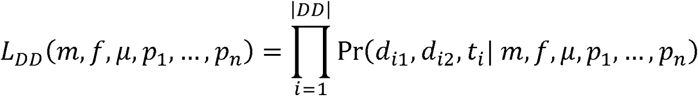

Where *d_i_*_1_ and *d_i_*_2_ are the test results of first and second visit of the women *i* in *DD*.

The full likelihood function is defined as:

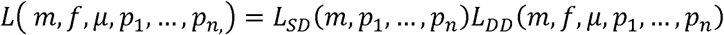

The explanation in detail for each situation can be seen in Figure S2.

### Fetal Fraction Modeling

The increase in fetal fraction from the 9th to the 17th week is modeled as follows:

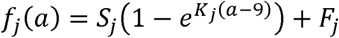

where *f_i_* represents the expected fetal fraction for group *j* (HBV-negative and HBV-positive under NIPT) at gestational age *a*. The parameter *K_j_* defines the growth rate, *F_j_* is the fetal fraction for group *j* at the 9th week, and *S_j_* represents the scale of the fetal fraction increase from the 9^th^ to the 17^th^ week. We assume that the residuals have Gaussian distribution:

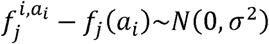

Where 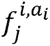 is the observed fetal fraction of the women *i* with gestational age *a_i_* and HBV-status *j*. The likelihood function is given by:

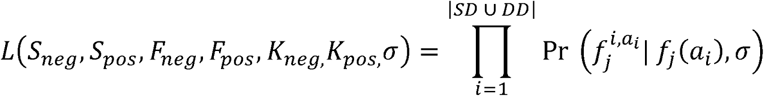

### Optimization

To enhance computational efficiency in the age-specific prevalence model, Bayesian estimation was employed as an alternative to maximum likelihood estimation. Specifically, the Metropolis algorithm was used to draw samples from the posterior distribution, with a uniform prior distribution [0,1] assumed for all parameters. Bayesian estimates were derived by calculating the mean of the posterior distribution.

In the fetal fraction model, we employed the Nelder-Mead algorithm for likelihood optimization. We statistically evaluated eight distinct epidemiological hypotheses using the Akaike Information Criterion (AIC), each varying in the degree to which the growth rate, scale parameter, and fetal fraction at the 9th week depend on HBV status. The detail of these eight hypotheses is depicted in Table S1. Confidence intervals for each parameter were calculated using likelihood profiling, focusing on the most plausible hypotheses.

### Statistical Analysis

Logistic regression was employed to explore the association between HBV infection and various covariates: number of visits, total sequencing reads, gestational age, fetal fraction, maternal age, and trisomy status. Additionally, a chi-squared test was performed to assess the consistency of HBV infection across the double NIPT visit cohort and to investigate the correlation between HBV cf-DNA levels and the outcomes of HBeAg and HBV-DNA tests. All data analyses and visualization were carried out using R (version 4.3.2) and Python (version 3.9).

## Result

### Data Characteristics

The trisomy rate throughout our dataset remains consistent at 3.4%, irrespective of the number of visits. Figure 1 provides a visual overview of key variables including maternal age, gestational age, total reads, and fetal fraction. In Panel A, the gestational age distribution is depicted, with an average of 11.71 weeks. The data skews rightward, with most women undergoing NIPT before 13 weeks, as reflected by the interquartile range of 10.14 to 12.57 weeks. Panel B displays the total number of reads, showing a unimodal distribution with a prominent peak around six million. While most samples are concentrated around this peak, there is a gradual tailing towards higher read counts, indicating some variability in sequencing depth. Finally, Panel C illustrates the distribution of fetal fractions of our dataset, peaking at around 12% and following a bell-shaped distribution. A minor proportion of samples show higher fetal fractions beyond 20%, as seen in the slight extension of the right tail. Further details on the cohort, particularly the comparison between women with one and two NIPT visits, can be found in Figures S3 and S4.

**Figure 1:**
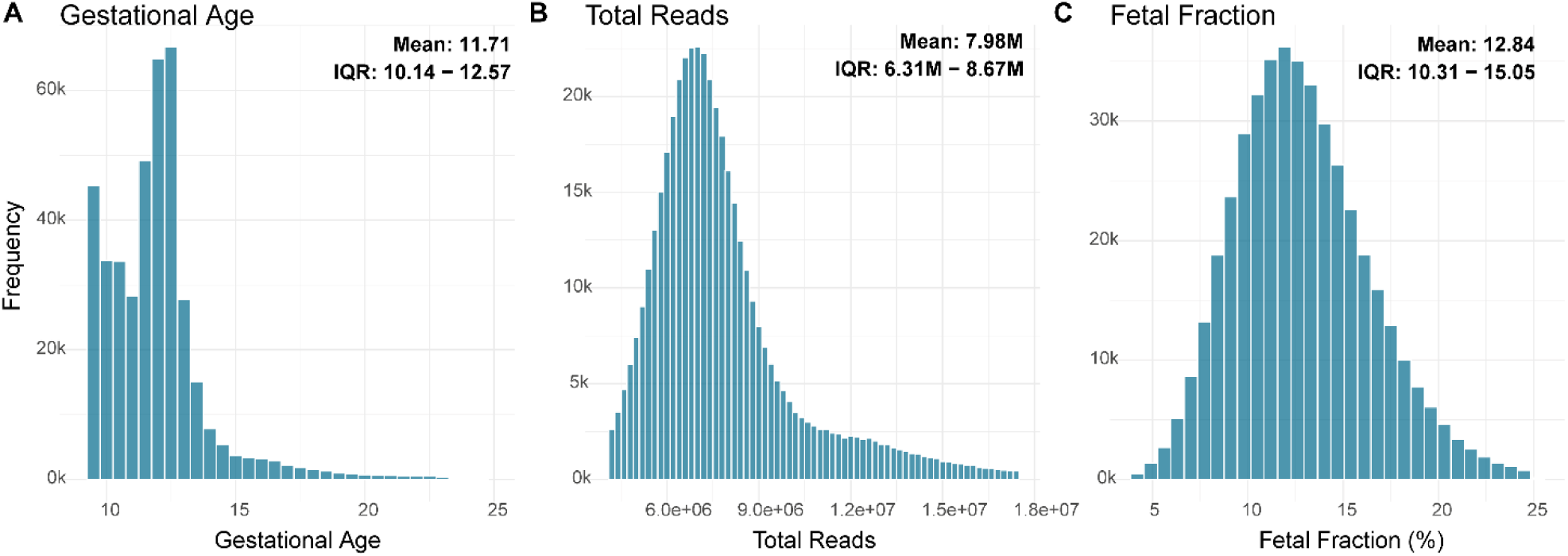
(A-C) Histograms - These histograms represent the distribution of gestational age, total reads, and fetal fraction across the entire dataset. Women observed twice due to multiple pregnancies are included twice in the dataset, each with a distinct profile.

### Association

To investigate factors linked to HBV infection, we conducted a logistic regression analysis using covariates collected during pregnancy visits and sequencing procedures: maternal age, gestational age, fetal fraction, total reads, and trisomy. We also included a binary variable, ‘two visits,’ indicating whether a woman was observed twice. The outcome variable was HBV status detected by NIPT at the first visit. As shown in Figure 2A, maternal age exhibits a strong association with HBV infection detected via NIPT (p = 6.89e-76). The odds of testing HBV-positive in an NIPT decrease to 95.3% of the previous odds with each additional year of maternal age. In other words, as maternal age increases, the likelihood of detecting HBV through the NIPT test decreases. In contrast, the number of visits and gestational age showed no significant association with HBV positivity under NIPT, whereas trisomy reached only marginal significance (p = 4.77 × 10⁻²). Although total reads were also significant (p = 5.76e-5), the odds ratio of 1.00000011 suggests a negligible effect on HBV testing outcomes. Importantly, our analysis revealed a significant negative association between fetal fraction and HBV infection (p = 3.33e-8). Given that fetal fraction may be less reliable in the absence of a Y chromosome ^37,38^, we further validated this finding by restricting the analysis to XY fetuses only (Figure S5). Consistent with the broader analysis, lower fetal fractions were associated with a higher likelihood of detecting HBV via NIPT. The odds ratios were 0.978 (95% CI: [0.971, 0.986]) for the full dataset and 0.979 (95% CI: [0.968, 0.989]) for the XY-restricted dataset.

**Figure 2:**
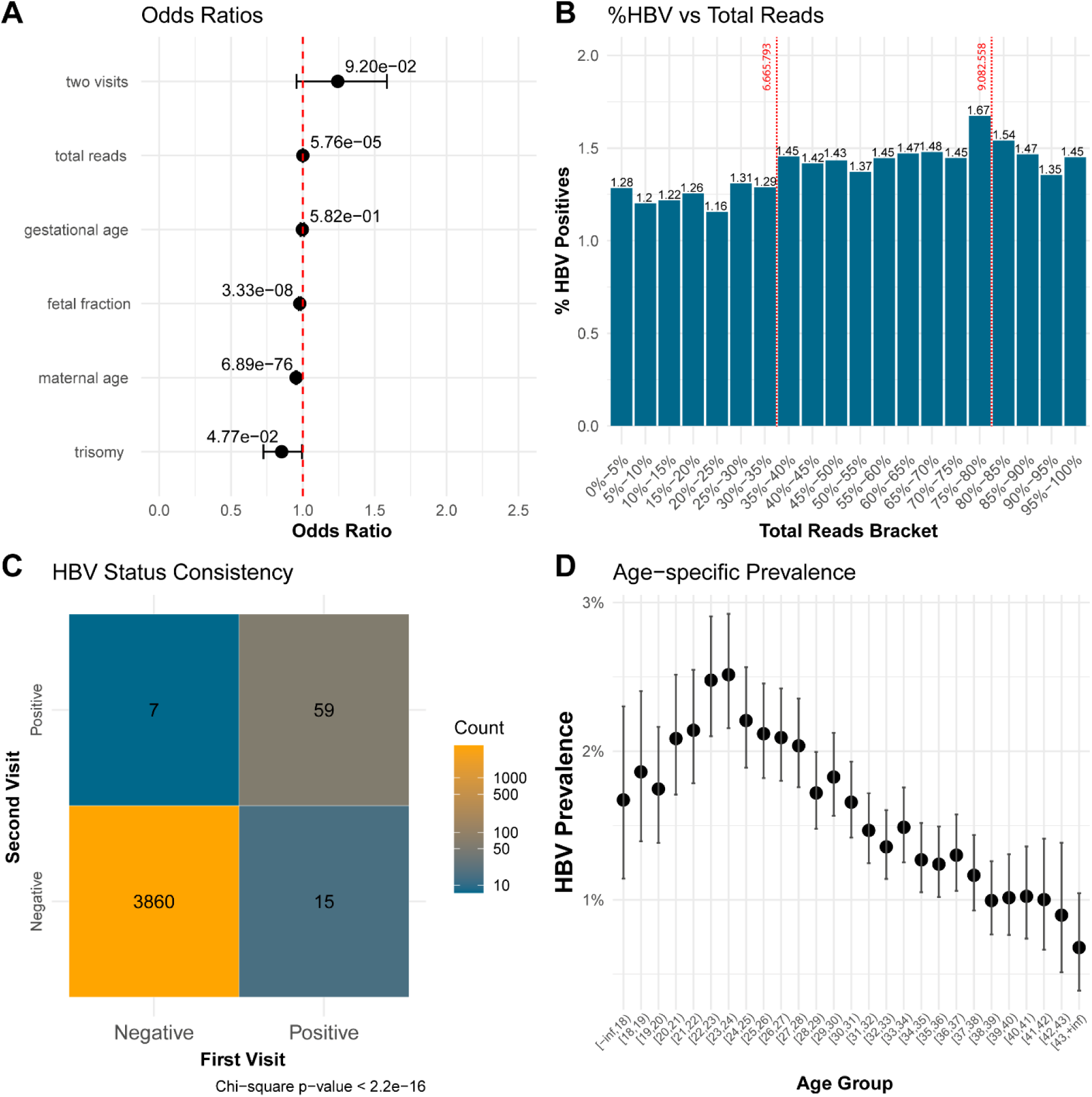
(A) Odd Ratios - A Forest plot illustrates the odds ratios for various factors associated with non-invasive prenatal testing (NIPT) outcomes. Odds ratios are shown with 95% confidence intervals, and the associated p-values are annotated on the plot. A red dashed line at an odds ratio of 1 serves as the reference baseline. The logistic regression model is based on outcomes from the first NIPT visit. (B) HBV Positivity and Total Reads - The bars show the proportion of HBV-positive cases observed. The red dashed lines represent the 50th and 80th percentile thresholds of total reads within the dataset, with corresponding values indicated. (C) HBV Consistency via NIPT - Consistency of HBV status between the first and second visits. The heatmap illustrates the number of women with consistent and inconsistent HBV status across both visits. The chi-square test result is displayed at the bottom of the panel. (D) Reconstructed Age-specific Prevalence - The dots and error bars show the Bayesian estimate of its 95% credibility intervals respectively.

### Fetal fraction

To investigate how fetal fraction relates to HBV status, we compared the growth patterns of fetal fraction between HBV-negative and HBV-positive groups during gestation from the 9th to 17th weeks. The best hypothesis which is selected via AIC revealed a key insight: while the growth rate and scale of fetal fraction were consistent across both groups, a significant difference in fetal fraction levels was already present at the earliest gestational age we observed (week 9) (Figure S9 and Table S1). Because the selected model shares the growth-rate and scale parameters across groups, the two fitted curves differ only by a constant offset within the observed window of weeks 9–17; we therefore report a stable association across this window and make no claim about gestation before week 9 or beyond week 17, which we did not observe.

To evaluate the fit of our model, we divided the data into 0.5-week intervals by gestational age and computed the average fetal fraction within each bin. These bin averages play a role in the trend of real data. Although the model itself lacks direct biological interpretation, it provides an excellent fit for the dataset. As seen in Figure 3, both the blue (HBV-negative) and red (HBV-positive) curves, which were derived using identical growth rate and scale parameters, align closely with the observed means for each gestational age bin, represented by the colored squares.

**Figure 3:**
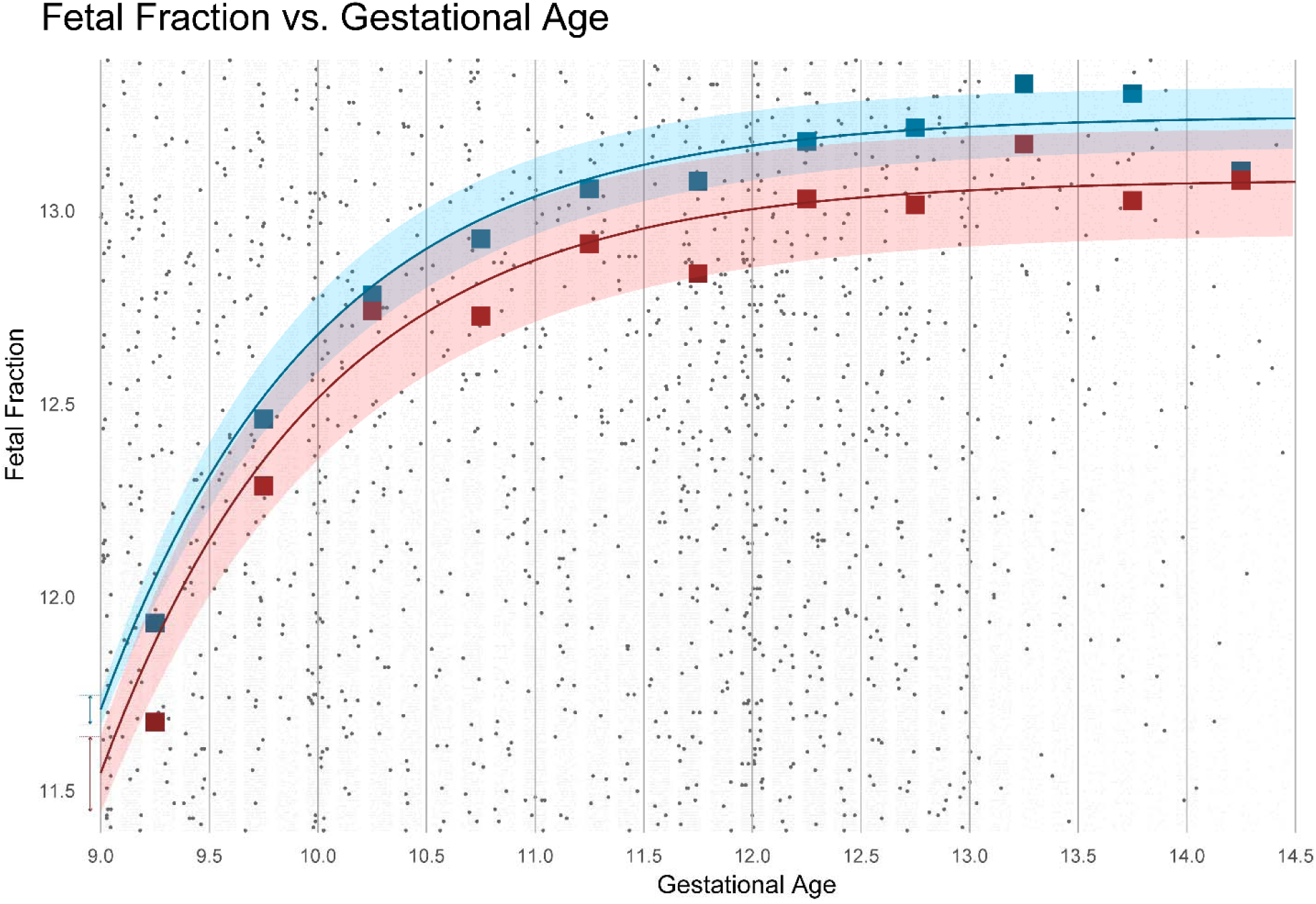
Relationship between fetal fraction and gestational age - Light and dark gray dots represent individual fetal fraction measurements from HBV-negative and HBV-positive participants, respectively. The blue and red curves depict the estimated relationship between fetal fraction and gestational age, with shaded areas indicating 95% confidence intervals (CI). Square markers represent the mean fetal fraction within each gestational age bin. The square boxes at [14, 14.5) reflect the mean fetal fraction for individuals with gestational ages between 14 and 17 weeks. Two-line segments on the x-axis display the 95% CI for fetal fraction at 9 weeks for HBV-negative and HBV-positive individuals.

The dynamics of fetal fraction in the graph reveal key insights into the differences between HBV-positive and HBV-negative pregnancies. At the 9th week of gestation, a noticeable disparity emerges: HBV-positive pregnancies have a fetal fraction of about 11.56% (95% CI: [11.45% - 11.65%]), while HBV-negative pregnancies show a slightly higher value, approximately 11.72% (95% CI: [11.68% - 11.76%]). This difference is statistically significant and persists during the early weeks of pregnancy. As gestational age progresses, the fetal fraction steadily increases in both groups. By the 12th week, the fetal fraction in both HBV-positive and HBV-negative pregnancies appears to plateau. This suggests that, despite an early discrepancy, the growth patterns of fetal fraction are similar beyond the 9th week. This indicates that the significant difference in fetal fraction at the 9th week is likely the main factor driving the observed overall differences between the two groups.

### HBV positivity and total reads

To examine the relationship between number of reads and HBV detection, we divided the dataset into 20 bins, each representing 5% quantiles of total reads distribution. Within each quantile, we calculated the percentage of HBV-positive samples. As illustrated in Figure 2B, HBV positivity rates rise significantly from 1.28% in the lowest bin to 1.67% in the bin representing the 75^th^ to the 80^th^ percentile (9.0M total reads). This upward trend suggests that increased sequencing depth initially enhances the sensitivity of HBV detection.

However, beyond this point, the HBV-positive rate drops slightly to 1.45%, indicating that further increases in sequencing reads do not yield substantial improvements in detection accuracy. Notably, the positivity rates observed between the 35^th^ and 70^th^ percentiles are comparable to those beyond the 85^th^ percentile, suggesting that once around 6.5M total reads is achieved, additional reads do not meaningfully enhance HBV detection via NIPT.

### HBV persistence in double visit data

NIPT-based HBV status was highly concordant between paired visits (Figure 2C). Of 3,941 individuals with two NIPT visits (mean interval 1.67 years, a subset exceeding 2.5 years), 3,860 were negative and 59 positive at both time points, whereas only 22 were discordant (15 positive-to-negative, 7 negative-to-positive) — an overall agreement of 99.4% (Cohen’s κ = 0.84; χ² p < 2.2 × 10⁻¹⁶). Because the interval spanned years, part of the discordance likely reflects genuine change in HBV status rather than assay error, so this represents a conservative bound on test variability. These paired-visit data, previously unavailable, indicate that HBV detection by NIPT is reproducible over time.

### Age-specific prevalence

The age-specific prevalence of HBV detected via NIPT was estimated using a Bayesian framework, employing Markov Chain Monte Carlo (MCMC) simulation with four million iterations. The model achieved an acceptance rate of 25.45%, and parameter convergence was confirmed through trace plots, autocorrelation analysis, and the Gelman-Rubin potential scale reduction factor (Figures S6 and S7, and Table S2). Figure 2D illustrates the reconstructed age-specific HBV prevalence, sampled from the posterior distribution (Figure S8), revealing considerable variation across age groups.

After accounting for factors such as infection dynamics, clearance rates, and detection uncertainty, HBV prevalence peaks at nearly 2.51% (95% Credible Interval: [2.15% - 2.92%]) in the 23-24 age group and remains elevated among women aged 22-28. Beyond the age of 28, however, there is a noticeable decline in prevalence, with rates progressively decreasing among older age groups. By age 43 and older, the prevalence drops to 0.68% (95% Credible Interval: [0.39% - 1.04%]). These results indicate that younger women, especially those in their early to mid-20s, are at a higher risk of HBV detection through NIPT, while the risk maybe decreases in women over 28.

### NIPT aligns with clinical markers of HBV infectivity

NIPT-derived HBV status aligned closely with clinical markers of viral activity. HBV-DNA titers were concordant with NIPT calls (Fig. 4a): NIPT-positive women almost invariably carried HBV-DNA above the threshold indicating maternal antiviral prophylaxis, whereas NIPT-negative women clustered at markedly lower titers. A single NIPT-positive woman fell below the treatment threshold; this case reflected an intrinsic discordance between her HBV-DNA and HBeAg results rather than an NIPT error (Table S3). Benchmarking NIPT against the composite of HBV-DNA and HBeAg status (Fig. 4b) confirmed a significant association (p = 7.20 × 10⁻⁷). Only two women were NIPT-negative yet positive by the composite measure, and both were among the individuals showing HBeAg–HBV-DNA discordance (Table S3) — indicating that the few mismatches trace to disagreement between the reference assays themselves rather than to NIPT.

**Figure 4:**
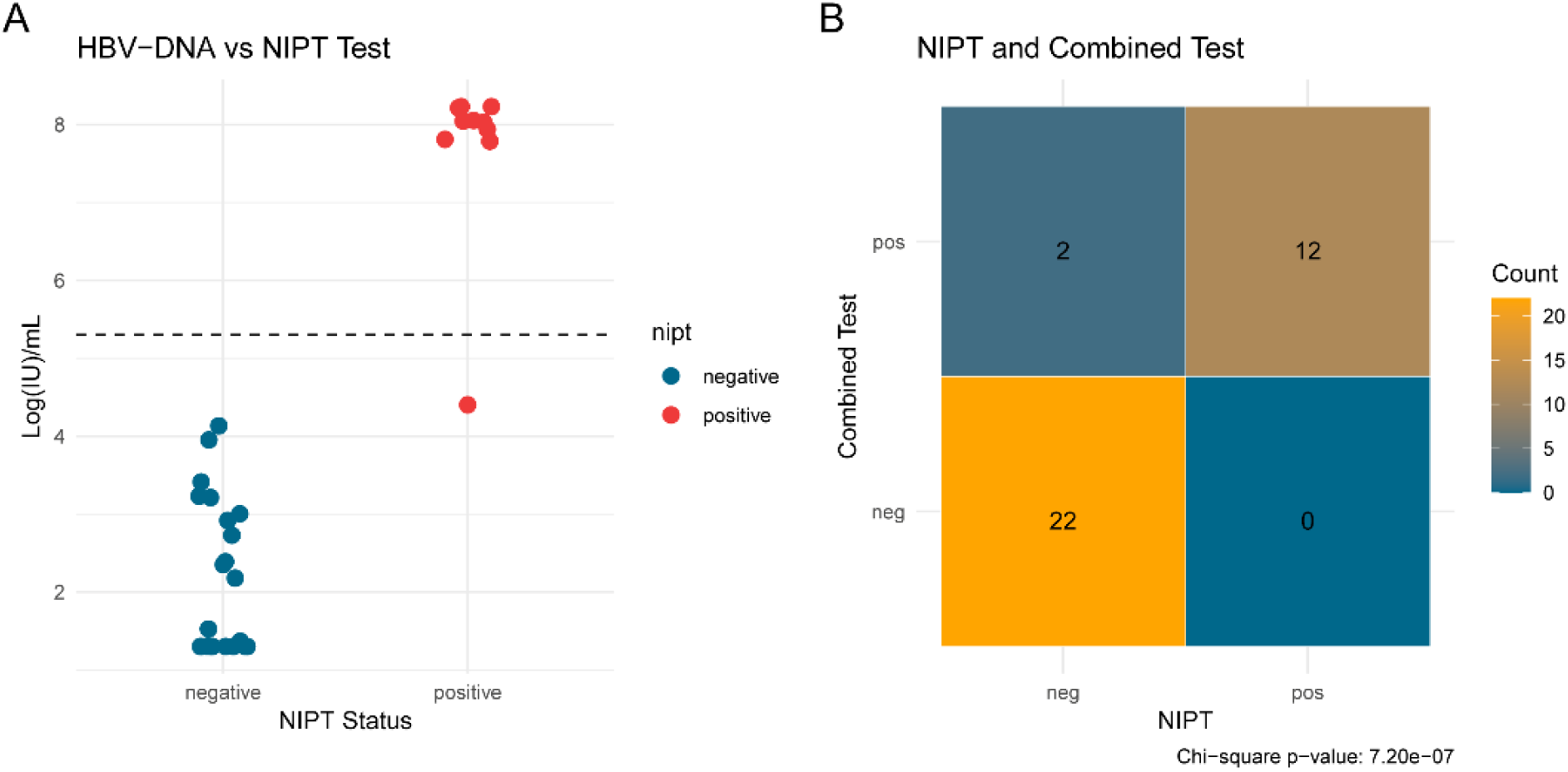
(A) Association between HBV-DNA and NIPT test results – The blue and red points represent the HBV-DNA levels in pregnant women with NIPT-negative and NIPT-positive results, respectively. The dashed line indicates the recommended threshold for initiating antiviral therapy. (B) Validation - This panel compares NIPT results with combined HBeAg and HBV-DNA testing (combined test).

## Discussion

In order to interpret the single and double NIPT visit datasets, the statistical model presented in this paper accounts for the HBV force of infection, clearance rate, and false detection. Although the age-specific prevalence generated by this approach demonstrates a clear trend with the peak can be observed in age class 23 at 2.5% (95%CI: [2.2%-2.8%]) (Figure 2D), this number is notably lower than established epidemiological data for high-endemicity countries. According to ^13–18^, HBV prevalence in the general population in Vietnam ranges from 8% to 20%, with approximately 10% among pregnant women ^19,20^. In our validation dataset, the prevalence of HBV under HBsAg test and NIPT test is 6.9% (95%CI:[5.0%, 9.2%]) and 2.1% (95%CI:[1.0%, 3.6%]) respectively. While this discrepancy might be attributed to shallow sequencing depth ^39^, our double-visit data and sensitivity analysis (Figure 2B and Figure 2C) indicate that even a fivefold increase in the number of reads is unlikely to resolve the issue. Hence, the disparity between the HBV prevalence computed from NIPT tests and HBsAg tests implies that the majority of HBsAg-positive women may have very low levels of HBV cfDNA, or possibly none at all.

The Hepatitis B virus (HBV) predominantly targets the liver, where it replicates within hepatocytes and subsequently releases viral particles into the bloodstream ^40–42^. These circulating particles comprise the complete virus, viral proteins, and genetic material. In the context of pregnancy, in addition to HBV-infected germline cells^43^, mother-to-child transmission can occur via two primary pathways: transplacental transmission and perinatal transmission, both of which rely on blood as the transmission vector ^44–46^. Therefore, the presence of HBV particles in the blood serves as a critical indicator of viral replication and infectivity. This explains why our NIPT findings show a stronger correlation with HBV-DNA and HBeAg tests, as these markers more accurately reflect viral load and active infection compared to HBsAg (Figure 2D, Figure 4A).

The double NIPT visit dataset offers a unique opportunity to evaluate the consistency of HBV detection via NIPT across two pregnancies in the same women. Among those who tested positive in the first NIPT, 80% also tested positive in the second, indicating that women with high and low infectivity are likely to remain so over several years. These results suggest that NIPT-based HBV status is concordant with HBeAg and HBV-DNA testing performed at the same time, supporting its potential as a complementary marker of viral infectivity.

Using big data, our analysis for the first time identifies an association between low fetal fraction and HBV infection detected via NIPT, addressing a significant gap in our understanding of HBV epidemiology during pregnancy. This discovery is contextualized by existing literature, which highlights two critical points. First, lower fetal fractions have been linked to reduced birth weight ^47^. Second, mothers with acute HBV infection are known to have a higher likelihood of delivering infants with lower birth weights ^13,48,49^. These connections raise the possibility that HBV status is linked to the placental contribution of cell-free DNA as early as the first trimester, potentially including cases of high viral replication and maternal infectivity. We emphasize, however, that this remains an association. The effect is small in magnitude (≈0.16 percentage points at week 9) and, although statistically robust given the cohort size, our fetal-fraction model was not adjusted for maternal age or body-mass index, both established determinants of fetal fraction ^50,51^. Notably, however, fetal fraction tends to be higher in younger and lower-BMI women, whereas HBV-positive women in our cohort were younger; age- and BMI-mediated confounding would therefore be expected to bias this estimate toward the null, making the observed reduction likely conservative rather than spurious. A direct association between HBV status and maternal BMI, independent of maternal age, cannot be excluded, as BMI was not available in our dataset. Our data also do not directly link low first-trimester fetal fraction to chronic placental inflammation ^52^; our data are consistent with an early association between HBV status and reduced fetal fraction within the first trimester, although how directly this association maps onto an effect of HBV on fetal or placental development remains unclear. This association is nonetheless concordant with reports of adverse fetal outcomes in HBV-positive mothers ^53^.

This study represents a pioneering effort to correlate HBV cell-free DNA with epidemiological patterns, leveraging a substantial dataset of 499,544 pregnant women from Vietnam, a region with high HBV prevalence. By integrating these findings with established epidemiological data, the research identifies HBV cf-DNA as a biomarker indicative of high viral infectivity. Notably, this research offers new insights into HBV epidemiology during pregnancy by revealing, for the first time at this scale, an early association between HBV status and reduced fetal fraction that is already detectable in the first trimester. From a public health perspective, NIPT could serve as a critical tool for identifying women with high HBV infectivity as early as the 12th week of pregnancy. By enabling early stratification of at-risk individuals, NIPT provides a valuable opportunity to implement targeted early interventions aimed at reducing mother-to-child transmission, one of the most significant pathways for HBV spread.

## Data Availability

All data supporting the findings of this study are available from the corresponding author upon reasonable request. Access to the sequencing data is subject to controlled access owing to patient privacy and institutional data-protection requirements.

## Author Contributions

VDN, PTN and MDP conceived and designed the study. MDP, HG, SHT, and HNN established and coordinated the collaboration. VDN, PTN, MFB and MDP performed the data analysis and drafted the manuscript. TTN and the collaborating investigators at the participating site carried out patient recruitment and data collection. All authors contributed to the interpretation of the results, critically revised the manuscript, and approved the final version for submission.

The lead author had full access to all study data and takes responsibility for the integrity of the data and the accuracy of the analysis.

## Funding

This study was funded by Gene Solutions, Vietnam. The funder provided support in the form of salaries for authors VDN, PTN, MDP, SHT, HNN, and HG, but did not have any additional role in the study design, data collection and analysis, decision to publish, or preparation of the manuscript. The specific roles of these authors are articulated in the author contributions section.

## Data Availability

The data that support the findings of this study are available from the corresponding author upon reasonable request.

## Competing Interests

VDN, PTN, MDP, SHT, HNN, MDP, and HG are employees of Gene Solutions, Vietnam. The other authors declare no competing interests.

## Ethics Statement

The study was approved by the institutional ethics committee of the University of Medicine and Pharmacy, Ho Chi Minh city, Vietnam. The study has followed the guidelines set by the University of Medicine and Pharmacy, Ho Chi Minh city, Vietnam, in handling human genetic data of the participants. The participants who performed NIPT triSure at Medical Genetics Institute, Vietnam, have approved and given written informed consent to the anonymous re-use of their genomic data for this study.

## Notes

### Competing Interest Statement

DNV, NTP, NHN, THS, PMD, and HG are employees of Gene Solutions, Vietnam. The remaining authors declare no competing interests. This affiliation did not influence the study design, data analysis, interpretation, or the decision to publish.

### Author Declarations

The Institutional Ethics Committee of the University of Medicine and Pharmacy at Ho Chi Minh City, Vietnam, gave ethical approval for this work.

