## Supplementary Tables and Figures for "Hepatitis B cell-free DNA in non-invasive prenatal testing as an early biomarker of viral infectivity"

**Supplementary Figures**

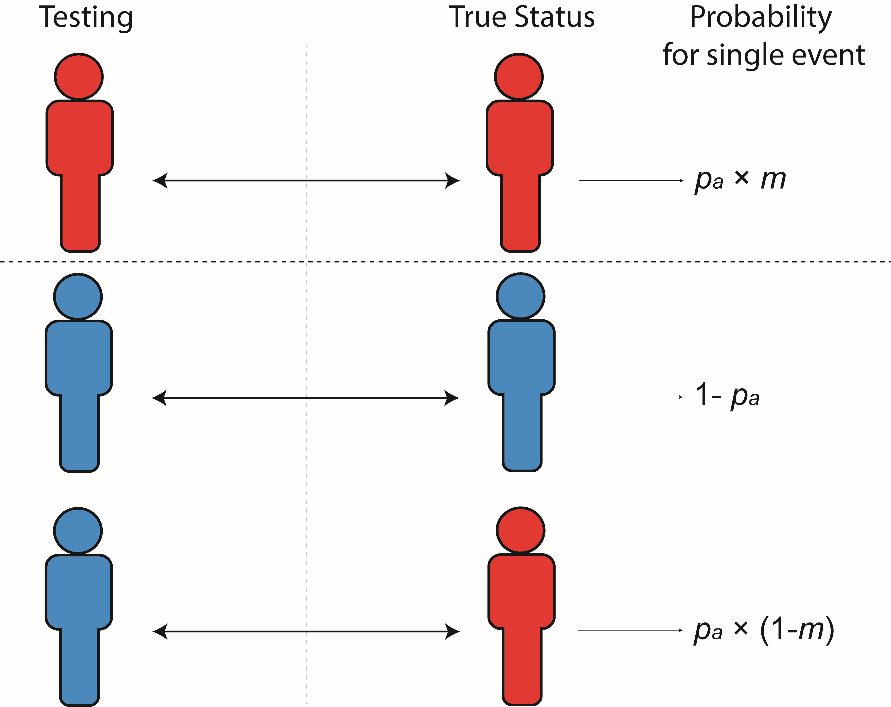

**Figure S1:** **Test Result and True Infection Status for Women with One NIPT Visit** - This diagram illustrates the relationship between NIPT test results and true infection status for pregnant women, with each row representing different scenarios. Red indicates a positive result or infection, and blue indicates a negative result or no infection. The final column details the calculation of the likelihood for each scenario. The notations used are consistent with those in the main text.

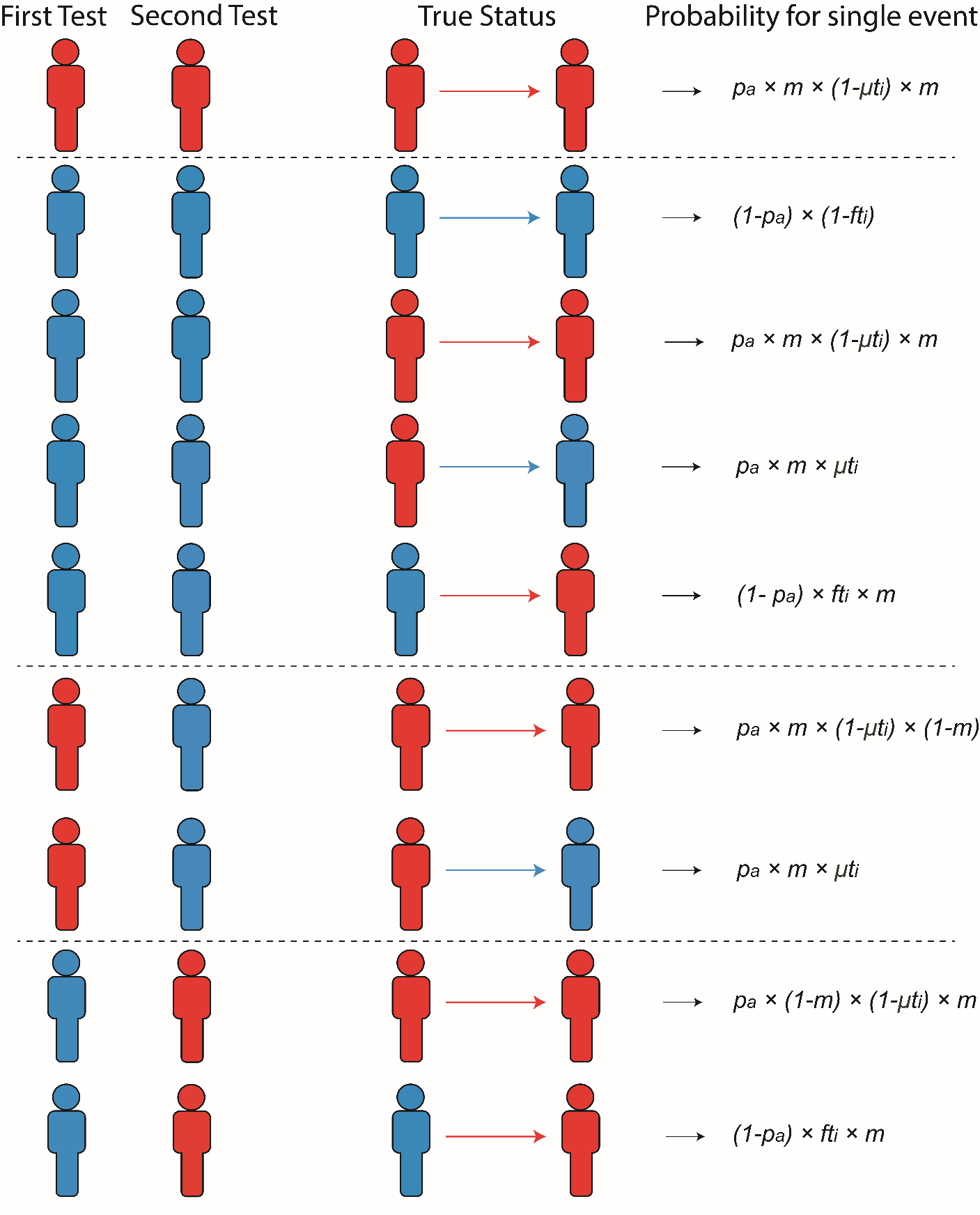

**Figure S2:** **Test Result and True Infection Status for Women with Two NIPT Visits** - This diagram illustrates the relationship between NIPT test results and true infection status for pregnant women with two NIPT visits. The color scheme follows Figure S1 with red and blue representing positivity and negativity respectively. The arrows show how true status may change between two visits. The final column details how the likelihood for each scenario is calculated. The notations used are consistent with those in the main text.

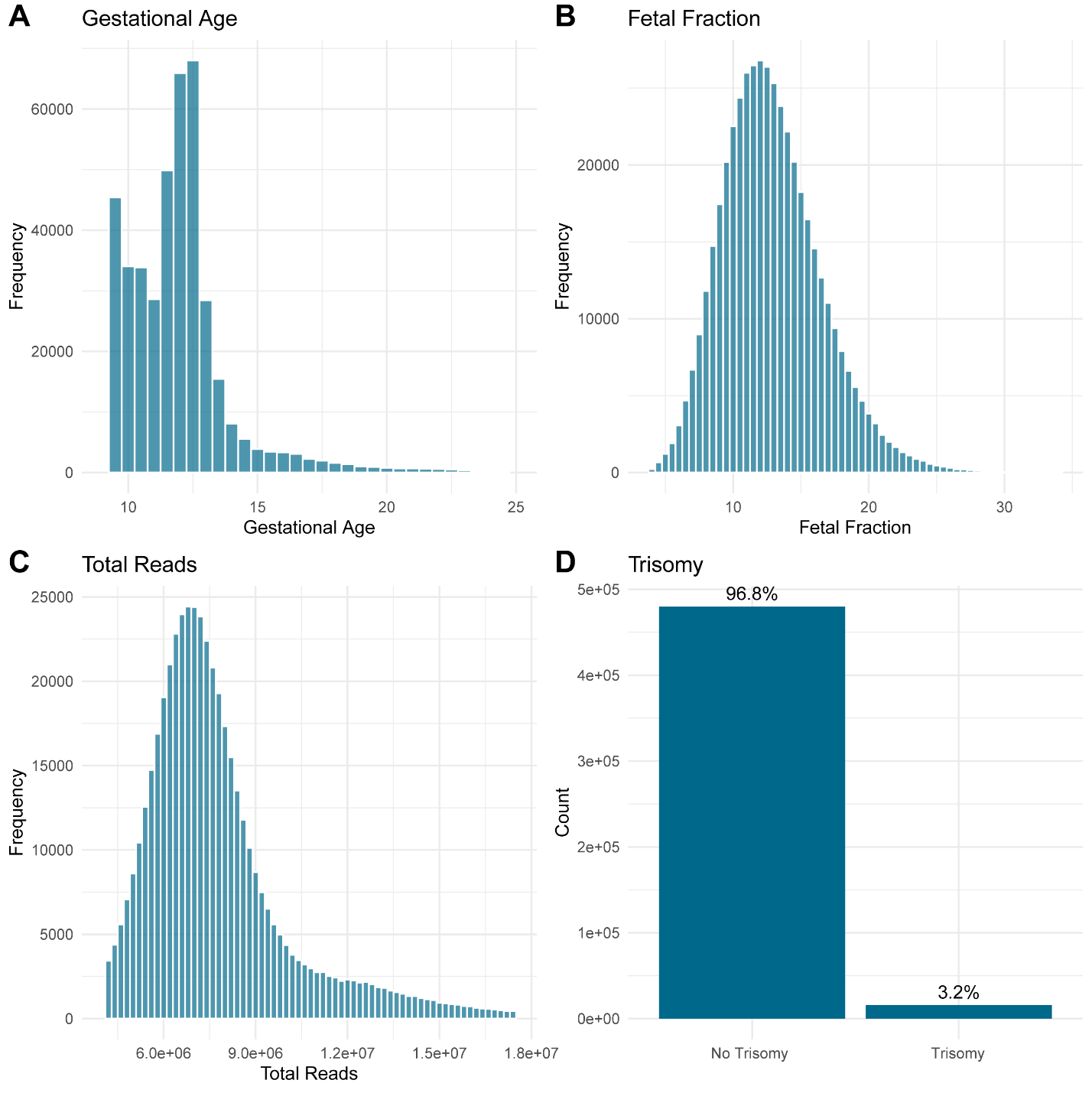

**Figure S3: Distribution of key variables of pregnant women with one NIPT visit**. **(A) Gestational Age** - Gestational age at the time of testing shows a peak around 11 weeks, with a range from 7 to 29 weeks. **(B) Fetal Fraction** - Fetal fraction is normally distributed with a peak around 12%. **(C) Total Reads** - The distribution of total reads is skewed with most samples around 2.5 million reads. **(D) Trisomy Status** - The trisomy status distribution reveals that 96.8% of the samples show no trisomy, while 3.2% are trisomy-positive cases.

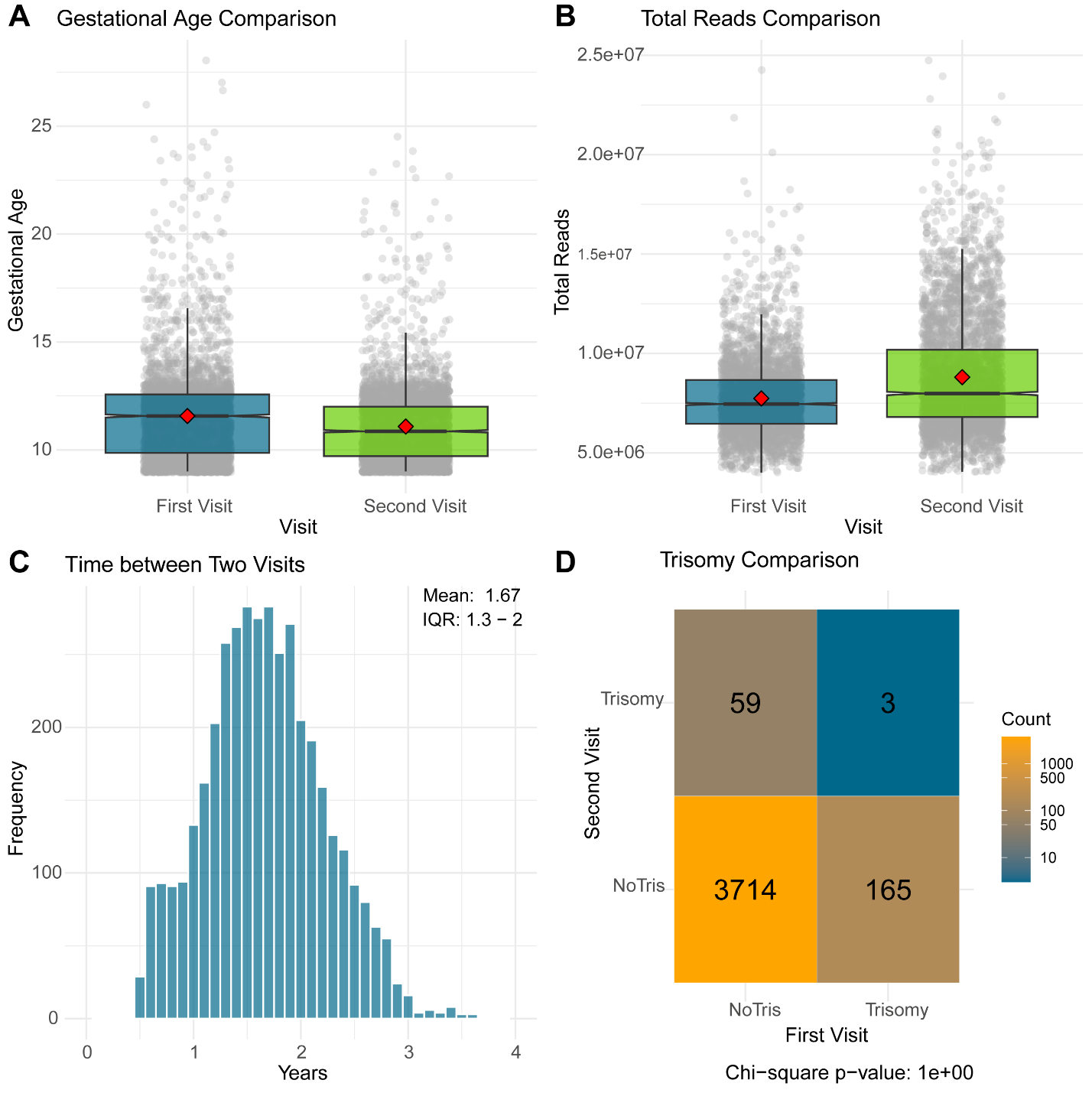

**Figure S4:** **Comparison of key variables between the first and second NIPT visits.** **(A) Gestational Age** - Gray dots represent individual data points, while the box plots summarize the gestational age distribution across the two NIPT visits, with medians indicated by red diamonds. A slight decrease in gestational age is observed at the second visit. **(B) Total Reads** - The format mirrors panel A. The comparison of total sequencing reads between the two visits shows consistent read numbers. **(C) Time Interval Between Visits** - The distribution of time between visits, with a mean of 1.67 years and an interquartile range from 1.30 to 2.00 years. **(D) Trisomy Comparison** - Analysis of trisomy status between the first and second visits, with a chi-square test indicating no significant difference in trisomy detection between the two visits.

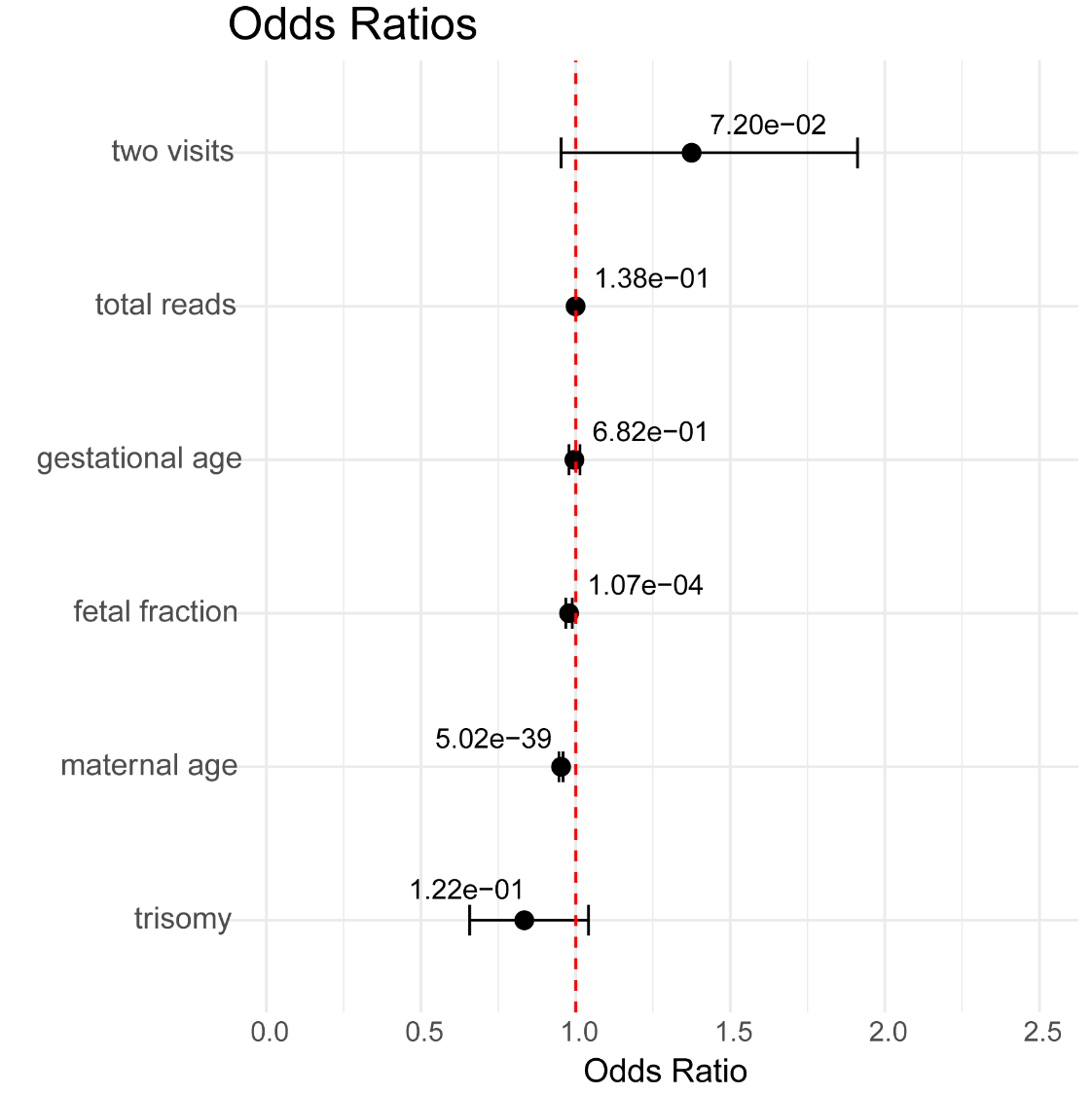

**Figure S5: Odds Ratios** - Forest plot illustrates the odds ratios for various factors associated with non-invasive prenatal testing (NIPT) outcomes. The “two visits” predictor is a binary variable indicating whether a woman underwent two NIPT visits. Odds ratios are shown with 95% confidence intervals, and the associated p-values are annotated on the plot. A red dashed line at an odds ratio of 1 serves as the reference baseline. The logistic regression model is based on outcomes from the first NIPT visit. This analysis focuses on fetuses with XY gender.

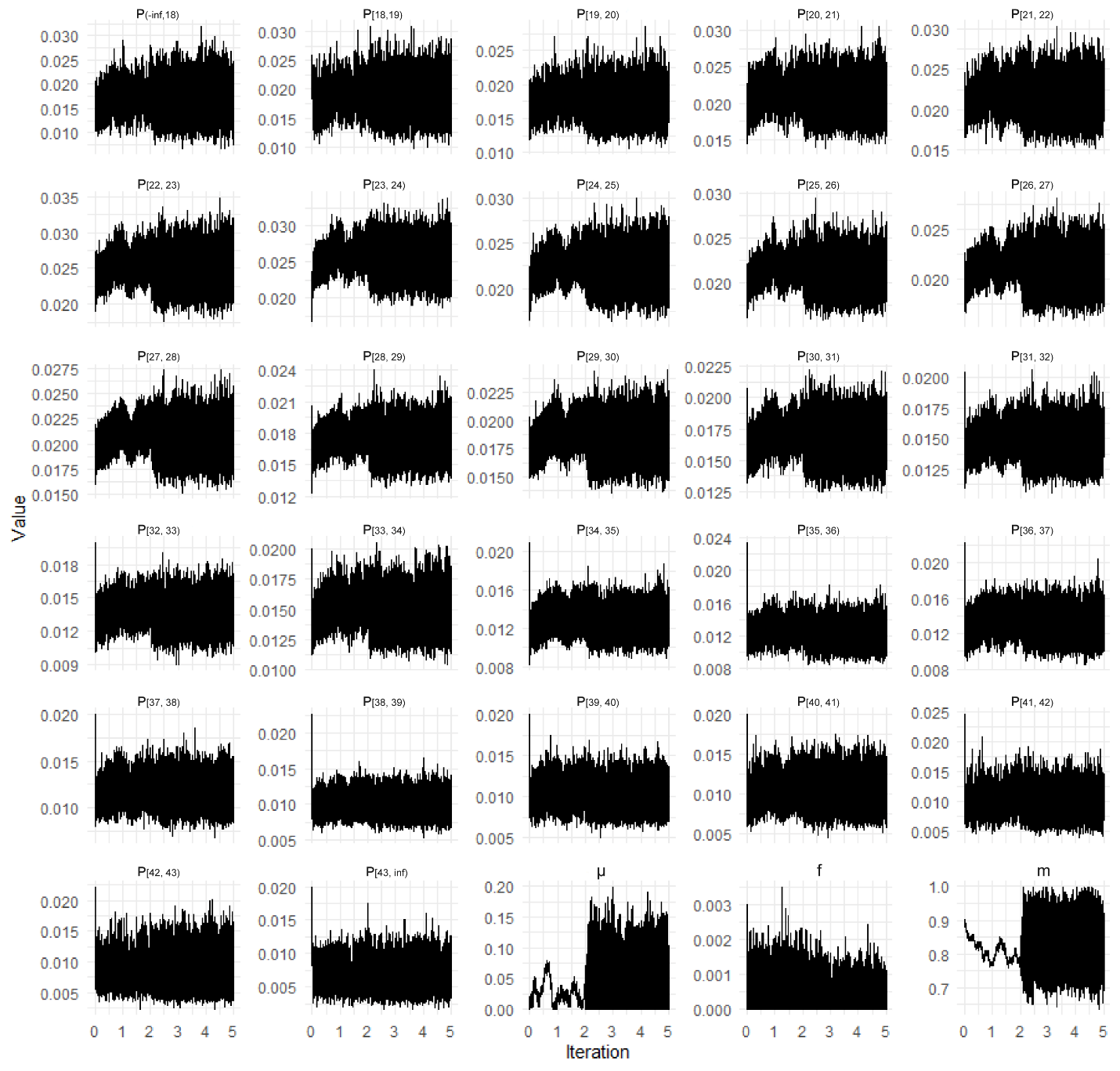

**Figure S6: Trace Plot** - The trace plots depict the MCMC chains for HBV prevalence across various age bins, along with key epidemiological parameters. The prevalence is denoted by P and age classes are shown in the subscript. The y-axis represents the parameter values, while the x-axis displays iterations in hundred thousand. A burn-in period of 200,000 iterations was applied, and only 500,000 first iterations are shown for clarity. The plots demonstrate the stability of the chains after the burn-in period, indicating sufficient mixing and convergence for most parameters.

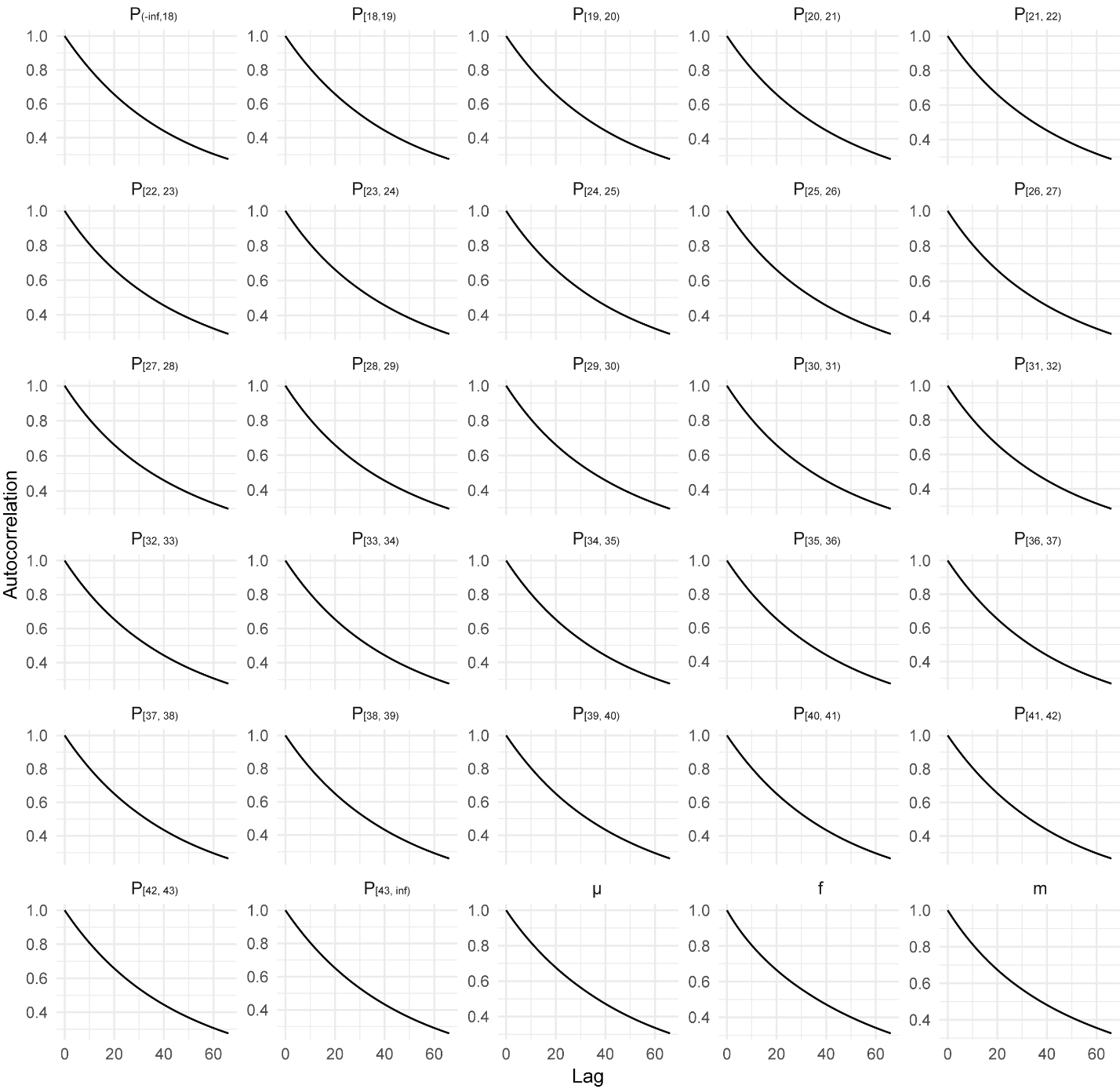

**Figure S7: Autocorrelation** - Autocorrelation plots for HBV prevalence across various age classes and key epidemiological parameters. Each panel shows the autocorrelation of the MCMC chains as a function of lag, with the y-axis representing the autocorrelation and the x-axis representing the lag. The steady decline in autocorrelation with increasing lag (consistently over parameters) suggests that the chains are well-mixed, indicating good convergence.

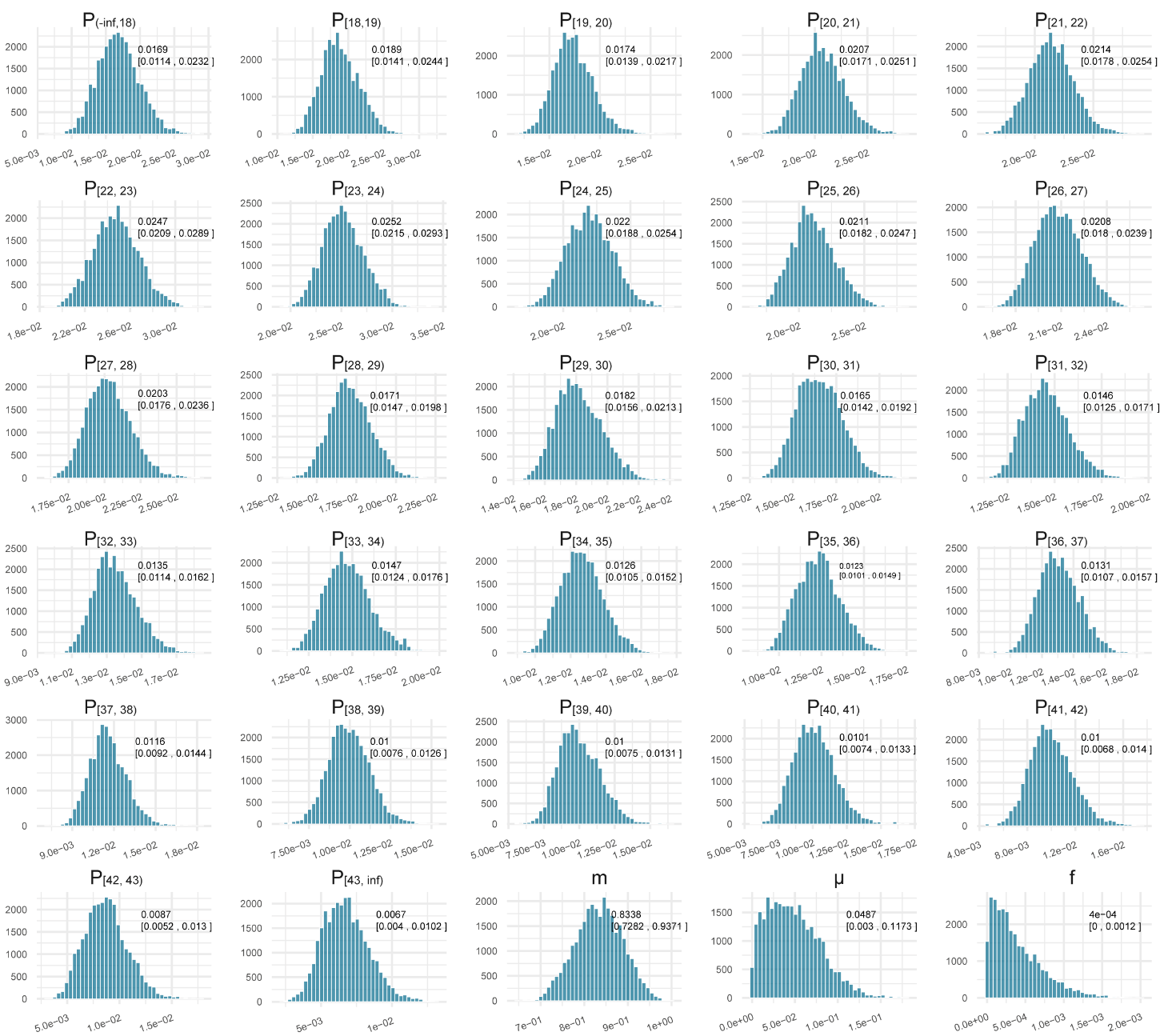

**Figure S8: Posterior distributions** - Histograms and 95% Credibility Intervals (CI) are generated using the last 30,000 states of the MCMC. The mean and 95% CI are displayed in the top right corner of each panel.

**Figure S9: Log-likelihood profile for the best hypothesis** - The horizontal red dashed line represents the 1.92 log-likelihood unit distance from the minimum value. Two vertical red dashed lines indicate the 95% confidence intervals for each parameter estimate.

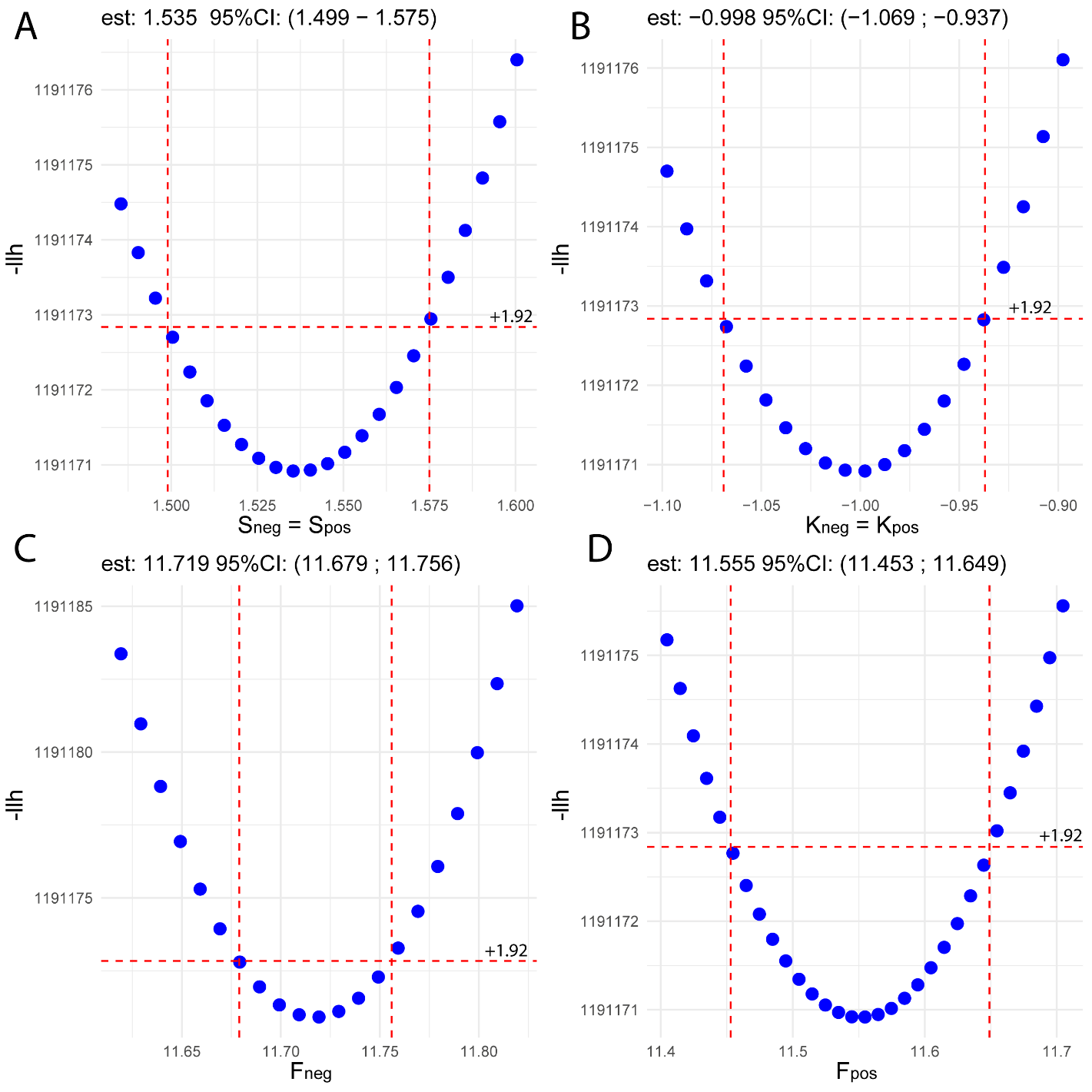

**Supplementary Tables**

**Table S1: AIC comparison of eight epidemiological hypotheses on fetal fraction increase** - The best-fitting hypothesis is highlighted in yellow, with constraints indicating the parameters considered HBV-independent in each case.

| Hypothesis | Constraints | #prms | -loglikelihood | AIC |
| --- | --- | --- | --- | --- |
| ff9_sc_sl | $S_{neg}=S_{pos}, F_{neg}= F_{pos},K_{neg}=K_{pos}$ | 4 | 1191177.099 | 2382362.197 |
| ff9_sl | $F_{neg}= F_{pos},K_{neg}=K_{pos}$ | 5 | 1191172.309 | 2382354.618 |
| sc_sl | $S_{neg}=S_{pos}, K_{neg}=K_{pos}$ | 5 | 1191170.915 | 2382351.829 |
| ff9_sc | $S_{neg}=S_{pos}, F_{neg}= F_{pos}$ | 5 | 1191172.568 | 2382355.136 |
| sl | $K_{neg}=K_{pos}$ | 6 | 1191170.778 | 2382353.556 |
| sc | $S_{neg}=S_{pos}$ | 6 | 1191170.918 | 2382353.835 |
| ff9 | $F_{neg}= F_{pos}$ | 6 | 1191172.044 | 2382356.088 |
| all | No constraint | 7 | 1191170.753 | 2382355.506 |

**Table S2: Gelman-Rubin Potential Scale Reduction Factor (PSRF)** - The PSRF was calculated for both univariate and multivariate analyses across various parameters in the HBV prevalence model. The multivariate PSRF, which evaluates the convergence of the entire set of parameters together, shows a value close to 1, indicating overall convergence. For the univariate PSRF, the table provides the point estimate and upper confidence interval (Upper CI) for each parameter. All values being close to 1 suggests good convergence for each individual parameter.

| **PRSF** | **parameter** | **Point estimate** | **Upper CI** | **Meaning in Model** |
| --- | --- | --- | --- | --- |
| Multivariate | NA | 1.00227 | NA | NA |
| Univariate | m | 1.000111 | 1.000378 | True Positive Rate |
|  | µ | 1.00005 | 1.000103 | Clearance Rate |
|  | f | 1.001066 | 1.002195 | Infection Force |
|  | p_(-inf,18)_ | 1.000131 | 1.000402 | HBV Prevalence for Age Group (-∞, 18) |
|  | p_[18, 19)_ | 1.000011 | 1.000031 | HBV Prevalence for Age Group [18, 19) |
|  | p_[19, 20)_ | 1.000318 | 1.001124 | HBV Prevalence for Age Group [19, 20) |
|  | p_[20, 21)_ | 1.000146 | 1.00031 | HBV Prevalence for Age Group [20, 21) |
|  | p_[21, 22)_ | 1.000075 | 1.000125 | HBV Prevalence for Age Group [21, 22) |
|  | p_[22, 23)_ | 1.000061 | 1.000087 | HBV Prevalence for Age Group [22, 23) |
|  | p_[23, 24)_ | 1.000267 | 1.00089 | HBV Prevalence for Age Group [23, 24) |
|  | p_[24, 25)_ | 1.00007 | 1.00026 | HBV Prevalence for Age Group [24, 25) |
|  | p_[25, 26)_ | 1.000124 | 1.000412 | HBV Prevalence for Age Group [25, 26) |
|  | p_[26, 27)_ | 1.000225 | 1.000779 | HBV Prevalence for Age Group [26, 27) |
|  | p_[27, 28)_ | 1.000407 | 1.001357 | HBV Prevalence for Age Group [27, 28) |
|  | p_[28, 29)_ | 1.000026 | 1.000055 | HBV Prevalence for Age Group [28, 29) |
|  | p_[29, 30)_ | 1.000027 | 1.000042 | HBV Prevalence for Age Group [29, 30) |
|  | p_[30, 31)_ | 1.000188 | 1.000512 | HBV Prevalence for Age Group [30, 31) |
|  | p_[31, 32)_ | 1.000211 | 1.000649 | HBV Prevalence for Age Group [31, 32) |
|  | p_[32, 33)_ | 1.000165 | 1.000542 | HBV Prevalence for Age Group [32, 33) |
|  | p_[33, 34)_ | 1.000165 | 1.000549 | HBV Prevalence for Age Group [33, 34) |
|  | p_[34, 35)_ | 1.000262 | 1.000913 | HBV Prevalence for Age Group [34, 35) |
|  | p_[35, 36)_ | 1.000309 | 1.001095 | HBV Prevalence for Age Group [35, 36) |
|  | p_[36, 37)_ | 1.000142 | 1.000454 | HBV Prevalence for Age Group [36, 37) |
|  | p_[37, 38)_ | 1.00015 | 1.000529 | HBV Prevalence for Age Group [37, 38) |
|  | p_[38, 39)_ | 1.000388 | 1.001363 | HBV Prevalence for Age Group [38, 39) |
|  | p_[39, 40)_ | 1.000128 | 1.000317 | HBV Prevalence for Age Group [39, 40) |
|  | p_[40, 41)_ | 1.00002 | 1.000058 | HBV Prevalence for Age Group [40, 41) |
|  | p_[41, 42)_ | 1.000189 | 1.000451 | HBV Prevalence for Age Group [41, 42) |
|  | p_[42, 43)_ | 1.000197 | 1.000673 | HBV Prevalence for Age Group [42, 43) |
|  | p_[43,inf)_ | 1.000136 | 1.000218 | HBV Prevalence for Age Group [43, +∞) |

**Table S3: Diagnosis data** - Diagnosis data collected from 40 pregnant women in our validation dataset (VD). Women who demonstrate the inconsistency between HBeAg and HBV-DNA test l are highlighted in yellow. The term 'NP' indicates 'not performed.' The term ‘CMB’ stands for ‘combined test’.

| **GA NIPT** | **NIPT** | **HBsAg** | **HBeAg** | **DNA** | **CMB** | **GA** |
| --- | --- | --- | --- | --- | --- | --- |
| 11.43 | 1 | 1 | NP | 1.64E+08 | pos | 11.43 |
| 13 | 1 | 1 | 1 | 1.08E+08 | pos | 26.29 |
| 19.29 | 1 | 1 | 1 | 1.70E+08 | pos | 19.29 |
| 12 | 1 | 1 | 1 | 6.46E+07 | pos | 13 |
| 11.57 | 1 | 1 | 1 | NP | pos | 11.57 |
| 11.71 | 1 | 1 | NP | 1.70E+08 | pos | 23.71 |
| 11.57 | 1 | 1 | 1 | NP | pos | 11.57 |
| 12 | 1 | 1 | 1 | 2.53E+04 | pos | 18 |
| 11.43 | 1 | 1 | NP | 1.11E+08 | pos | 15.43 |
| 11.43 | 1 | 1 | 1 | 1.13E+08 | pos | 12 |
| 12.43 | 1 | 1 | NP | 8.69E+07 | pos | 12.43 |
| 12.14 | 1 | 1 | NP | 6.12E+07 | pos | 16.14 |
| 12.43 | 0 | 1 | 0 | 3.36E+01 | neg | 6 |
| 12.71 | 0 | 1 | 0 | 2.25E+02 | neg | 25.71 |
| 12.43 | 0 | 1 | NP | NP | NP | NA |
| 12.14 | 0 | 1 | NP | 2.00E+01 | neg | 12.14 |
| 13.43 | 0 | 1 | NP | 1.36E+04 | neg | 13.43 |
| 11.57 | 0 | 1 | NP | 2.00E+01 | neg | 25.57 |
| 11.14 | 0 | 1 | NP | NP | NP | NA |
| 11.71 | 0 | 1 | NP | 1.51E+02 | neg | 23.71 |
| 12 | 0 | 1 | NP | 2.01E+01 | neg | 12 |
| 11.14 | 0 | 1 | 0 | 5.36E+02 | neg | 37.14 |
| 12.43 | 0 | 1 | NP | 1.01E+03 | neg | 27.43 |
| 15.29 | 0 | 1 | NP | NP | NP | NA |
| 11.86 | 0 | 1 | NP | 1.63E+03 | neg | 11.86 |
| 11.43 | 0 | 1 | 0 | 2.59E+03 | neg | 15 |
| 12.29 | 0 | 1 | NP | 2.00E+01 | neg | 25.29 |
| 11.71 | 0 | 1 | 0 | 8.30E+02 | neg | 16.29 |
| 12.29 | 0 | 1 | NP | 2.00E+01 | neg | 21.29 |
| 14.57 | 0 | 1 | 0 | 2.45E+02 | neg | 16 |
| 11.86 | 0 | 1 | 0 | 2.00E+01 | neg | 25 |
| 14.86 | 0 | 1 | NP | 2.00E+01 | neg | 26.71 |
| 13 | 0 | 1 | NP | NP | NP | NA |
| 12.14 | 0 | 1 | 1 | 8.98E+03 | pos | 7 |
| 12.29 | 0 | 1 | NP | 1.70E+03 | neg | 17.29 |
| 12.43 | 0 | 1 | 1 | 2.00E+01 | pos | 25.43 |
| 12.86 | 0 | 1 | 0 | 2.34E+01 | neg | 26.71 |
| 12.14 | 0 | 1 | NP | 2.00E+01 | neg | 12 |
| 11.43 | 0 | 1 | 0 | 2.00E+01 | neg | 16.43 |
| 12.43 | 0 | 1 | NP | 2.00E+01 | neg | 25.29 |
